# Symptom based and transmission-prevention based testing in long-term care facilities: Symptomatology, clinical course and mortality for residents with COVID-19

**DOI:** 10.1101/2020.10.28.20221275

**Authors:** Kelly C. Paap, Anouk M. van Loon, Sarian M. van Rijs, Esther Helmich, Bianca M. Buurman, Martin Smalbrugge, Cees M.P.M. Hertogh

**Affiliations:** Department of Medicine for Older People, Amsterdam Public Health Research Institute, Amsterdam University Medical Center, van der Boechorststraat 7, 1081 BT, Amsterdam, The Netherlands; Amsta Healthcare Organisation, Amsterdam, The Netherlands; Department of Internal Medicine, section of Geriatric Medicine, Amsterdam Public Health Research Institute, Amsterdam University Medical Center, van der Boechorststraat 7, 1081 BT, Amsterdam, The Netherlands

## Abstract

**Objectives:** Initially, for preventing COVID-19 transmission in long-term care facilities (LTCF) primarily rely on presence of core symptoms (fever, cough, dyspnea), but LTCF residents may also show an atypical course of a SARS-CoV-2 infection. We described the clinical presentation and course of COVID-19 in LTCF residents who were tested either because of presence of core symptoms (S-based) or because of transmission prevention (TP-based)

**Design:** Retrospective cohort study.

**Setting and participants:** Amsta (Amsterdam, The Netherlands), is a 1185-bed LTCF. All LTCF residents who underwent SARS-CoV-2 RT-PCR testing between March 16, 2020 and May 31, 2020 were included (n = 380).

**Measures:** Clinical symptoms, temperature and oxygen saturation were extracted from medical records, 7 days before testing up to 14 days after testing.

**Results:** SARS-CoV-2 was confirmed in 81 (21%) residents. Of these 81, 36 (44%) residents were tested S-based and 45 (56%) residents were tested TP-based. Yet, CT-values did not differ between the groups. In the 7 days prior to the test the most common symptoms in both groups were: falling (32%), somnolence (25%) and fatigue (21%). Two days before the test, we observed a stronger decrease in oxygen saturation and an increase in temperature for the S-based group compared to the T-based group that remained up to 10 days after testing. Residents with in the S-based group were 2.5 times more likely to decease within 30 days than residents in the TP-based group (HR, 2.56; 95% 1.3 to 5.2). Even though, 73% of the T-based group did eventually developed core symptoms.

**Conclusion and implications:** Many LTCF residents with a positive PCR did not have core symptoms when tested but had other signs/symptoms in the week before the positive test. Testing policies should therefore be adjusted to prevent transmission. Daily measures of temperature and oxygen saturation can contribute to earlier detection.

## Introduction

Residents in long-term care facilities (LTCFs) account for a disproportionate number of COVID-19 related deaths, making up half of all COVID-19 related deaths in Western countries^1^. This might be due to a fast and “invisible” transmission combined with the clinical frailty of the LTCF population who, in general, has severe medical comorbidities^2^. Older age and the presence of chronic medical comorbidities are associated with increased risk of mortality in the current pandemic^3^. The high prevalence of functional and cognitive impairment add to susceptibility of SARS-CoV-2 infections in LTCF residents^4^. Initially the case definition of COVID-19, and reason to do a Reverse Transcription Polymerase Chain Reaction (RT-PCR) test, was presence of at least one of the following core symptoms:fever of feverish feeling, cough or dyspnea^5^. Yet, the first major study describing COVID-19 in a LTCF showed that more than half of all the positive tested patients (56.5%) did not have these core symptoms when tested positive, but other symptoms such as sore throat, chills, malaise, and diarrhea^2^. From previous studies it is known that residents in LTCFs with infections frequently do not have fever^6-8^. Most of the LTCFs residents with COVID-19 also did not develop fever, but still had temperature elevations^9,^. If the focus for testing is only on residents with core symptoms, many residents with COVID-19 are missed, with consequences for transmission prevention.

Furthermore, as a large proportion of all LTCF residents has cognitive impairment and impaired ability to communicate it is often difficult to accurately observe and interpret symptoms. Therefore, although residents may only show a rise of temperature and not fever, measuring objective parameters like temperature and oxygen saturation may provide more reliable information in the LTCF population in addition to observing symptoms ^10-12^.

This study, in which LTCF residents were tested either because of presence of core symptoms (symptom based testing) or because of transmission prevention (transmission-prevention based testing), aims to describe the clinical presentation and course of COVID-19 (including 30-day mortality) in both groups and differences between both groups. A second aim is to compare the CT value of the RT-PCR test between the two groups.

## Methods

### Study population

This observational study was conducted in residents living in the LTCF Amsta (Amsterdam, The Netherlands), a 1185-bed skilled nursing facility with 18 locations spread over Amsterdam.

### Study design

In this retrospective cohort study, all residents who underwent a Reverse Transcription Polymerase Chain Reaction (RT-PCR) test from March 16, 2020 (the day we started testing), up to May 31, 2020 were included.

RT-PCR tests were performed with a throat and nasopharyngeal swab. Patients who had a positive RT-PCR test were considered to be COVID-19 confirmed (cycle threshold (CT) value was also reported). A test was performed a) when a resident had core symptoms according to Dutch guidelines (fever, cough or dyspnea) ^5^ (referred to as symptom based testing (S-based)) for transmission prevention (referred to as transmission-prevention based testing (TP-based)) if a resident had potentially contacted with individuals with confirmed COVID-19 (residents as well as healthcare workers), after hospital admission or outpatient clinic visit or when residents were newly admitted to the LTCF. Residents on some wards were tested several times because one of the residents turned out to be positive. It sometimes happened that residents with already confirmed COVID-19 were tested again on these wards.

### Data collection

Data about age, gender, BMI, type of ward, comorbidities and renal function of all residents that were tested on COVID-19 were extracted from the electronic health record (EHR) PinkRoccade Healthcare myCaress (myCaress).

For all residents with a confirmed COVID-19, we searched for clinical symptoms and for temperature and oxygen saturation data in the EHR from 7 days before testing up to 14 days after testing. The oxygen saturation was measured with a pulse oximeter and the body temperature was measured differently per ward, rectally or tympanum. Clinical symptoms were extracted by reviewing the EHR, using registrations of physicians as well as of nursing staff. We searched for clinical symptoms that were new or significantly changed (i.e. a change from incidental to more frequent falling) for each individual resident. A symptom was regarded as new if it had not been reported in the previous 4 weeks, and changed if it significantly changed compared to the previous 4 weeks. From the day that the first patient tested positive, it was agreed for the whole LTCF Amsta to measure the temperature and the oxygen saturation daily in all residents and to register it in the EMR (specific sections). Data about temperature and oxygen saturation were extracted from this specific sections. Data about 30-day mortality were also collected from the EHR.

### Statistical analysis

Baseline characteristics of all tested residents were analysed descriptively. Clinical symptoms, temperature and oxygen saturation of residents with confirmed COVID-19 were also analysed descriptively. To indicate the number of missing values, the N on which percentages were calculated, are reported. All continuous data were presented as means with standard deviation and 95% confidence intervals (CI). To evaluate the differences in clinical symptoms, temperature and oxygen saturation between the two groups of residents with confirmed COVID-19 (symptom based testing vs transmission-prevention based testing) group ANOVA’s were applied. We considered a P-value of <.05 to be statistically significant. Survival curves on 30-day mortality were estimated based on the days between the positive RT-PCR test and the date of death using Kaplan Meier curves in residents with confirmed COVID-19 (symptom based testing and transmission-prevention based testing). The mortality rate of residents with confirmed COVID-19 (symptom based testing vs transmission-prevention based testing) was compared using a Cox proportional hazard model, using three models. Model 1 unadjusted, model 2 included gender and age, model 3 included gender, age and reduced renal function. Reduced renal function was significant in the univariate analysis and was much more common in the S-based group. Results are presented with 95% confidence intervals and all reported P values are two-sided.

Finally, using ANOVA, we compared the CT-value of both groups (symptom based testing vs transmission-prevention based testing).

All analyses were performed with the use of the SPSS statistical package, version 26.0 (SPSS, Armonk, NY: IBM Corp).

### Ethics

The Medical Ethics Committee of the Academic Medical Centre in Amsterdam approved the study protocol.

## Results

On March 16, 2020, the date the first RT-PCR SARS-CoV-2 test took place, Amsta had a bed capacity for 1185 residents. Between March 16th and May 31th, 2020, 380 residents underwent a RT-PCR test for COVID-19 (601 tests). COVID-19 was ruled out in 299 (79%) residents and confirmed in 81 (21%) of residents. Of these 81 residents, 36 (44%) residents were tested because of presence of core symptoms (S based), compared to 45 (56%) residents who tested positively but were tested because of transmission prevention (TP based). (Figure 1)

### Patient characteristics

Mean age for the COVID-19 residents was 80 years (SD ± 11.2) in the S-based group and 82 years (SD ± 8.7) in the TP-based group. There were more women in both groups, S-based (53%) and TP-based (60%). In residents who were tested but for whom COVID-19 was ruled out the mean age was 79 years (SD ±11.1), and the majority were woman 62%. Almost all residents in the TP-based group had dementia (96%) whereas in the S-based group this was 83%. In residents tested with COVID-19 ruled out, just over half of these residents had dementia (59%). After dementia, cardiac problems, hypertension and a reduced kidney function were the most common comorbidities in residents who tested positive or negative for COVID. (Table 1.)

**Table 1.** Patient characteristics.

|  | Mean (SD), % (n) |  | COVID-19 ruled out |
| --- | --- | --- | --- |
|  | Confirmed COVID-19 (symptom based testing) | Confirmed COVID-19 (transmission-prevention based testing) |  |
| N | 44 (36) | 56 (45) | 79 (299) |
| Age, y | 80 (11.2) | 82 (8.7) | 79 (11.1) |
| **Sex** |  |  |  |
| Male | 47 (17) | 40 (18) | 38 (115) |
| Female | 53 (19) | 60 (27) | 62 (184) |
| BMI (kg/m<sup>2</sup>) | 24 (4.6) | 24 (5.1) | 24 (5.3) |
| **Ward type** |  |  |  |
| Psychogeriatric | 58 (21) | 91 (41) | 46 (136) |
| Somatic | 0 | 2 (1) | 22 (67) |
| Short term care | 11 (4) | 0 | 18 (53) |
| Geriatric psychiatry | 6 (2) | 0 | 9 (26) |
| Neuropsychiatry | 25 (9) | 7 (3) | 6 (17) |
| **Comorbidities** |  |  |  |
| Dementia | 83 (30) | 96 (43) | 59 (175) |
| Cardiac history | 31 (11) | 33 (15) | 42 (126) |
| Chronic respiratory disease | 11 (4) | 16 (7) | 18 (55) |
| Hypertension | 25 (9) | 24 (11) | 40 (120) |
| **Renal function** |  |  |  |
| Normal renal function (eGFR ≥ 60) | 28 (10) | 73 (33) | 51 (153) |
| Reduced renal function (eGFR < 60) | 64 (23) | 9 (4) | 40 (119) |
| No renal function registered | 11 (4) | 18 (8) | 9 (27) |

### CT-values

CT-values were available for 64/81 positive residents; the mean CT-value was 26.2 (range 12.5-40.4). Symptom based tested residents (n=27) had a median CT-value of 24.8 (range 15.6 -39), transmission-prevention based testing residents (n=37) had a median CT-value of 24.9 (range 12.5 -40.4). There were no significant differences between these groups F(1.63) = .011, P = .919.

### Symptoms 7 days before RT-PCR-confirmed SARS-CoV-2 infection

In the 7 days before a positively confirmed COVID-19 test, only fever was reported of the core symptoms and only in the S-based group. No core symptoms were present om the TP based group. The most common other symptoms were: falling, somnolence, fatigue and asthenia (Table 2). In the TP based group of patients falling occurred more often than in the S-based group (42% vs. 19%). Prior to the moment of testing other reported symptoms were: restlessness, dysphagia, diarrhea, sore throat, drowsiness and headache.

**Table 2.**
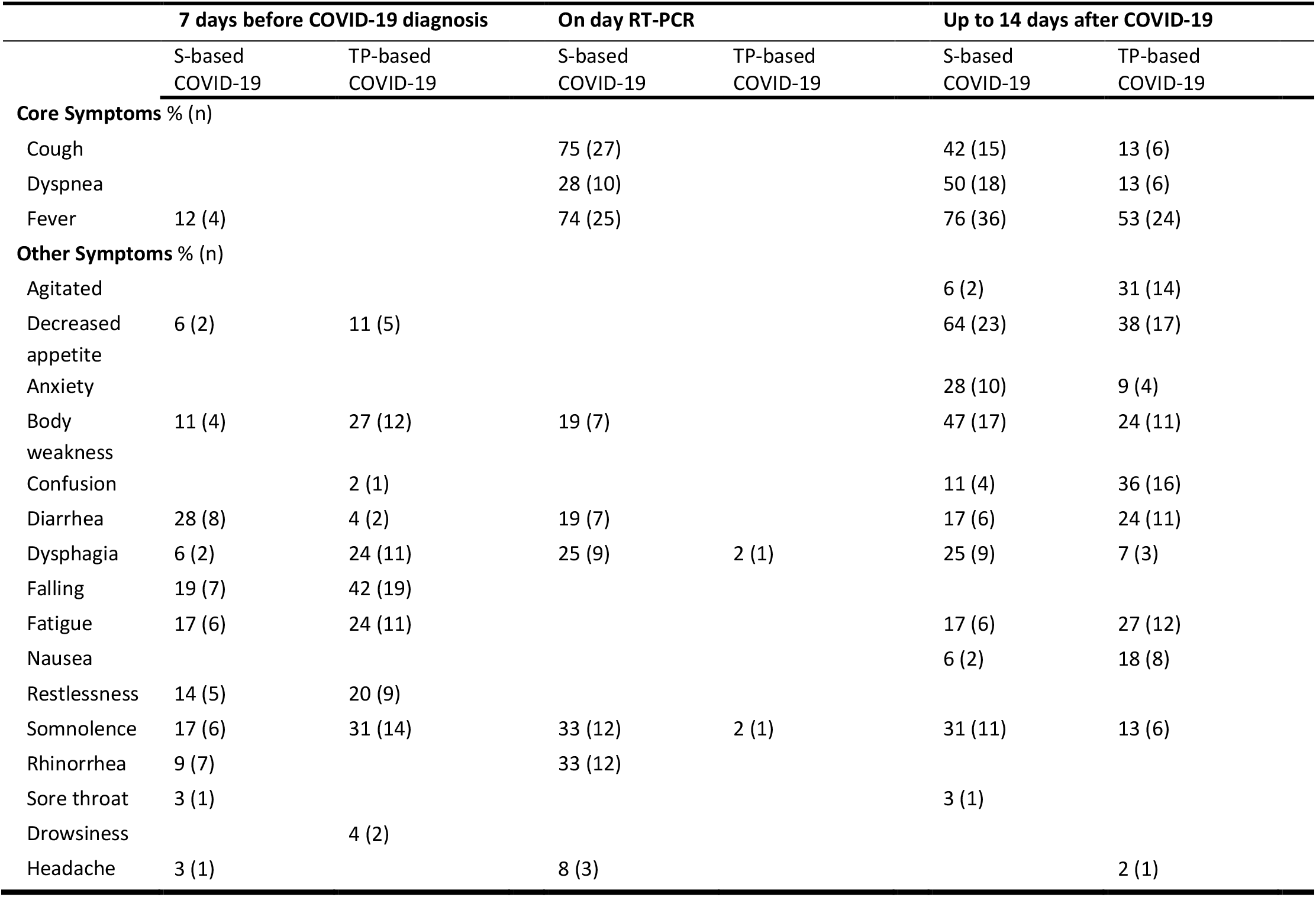
Core symptoms, other symptoms of COVID-19 confirmed residents.

### Signs/Symptoms on the day of the RT-PCR-test

On the day of COVID-19 testing, cough and fever were more frequent than dyspnea in the S-based group, 75%, 74% and 28% respectively. Other symptoms in the S-based group that were reported were somnolence (33%), rhinorrhea (33%), dysphagia (25%), body weakness (19%) and diarrhea (19%).

### Signs/Symptoms up to 14 days after RT-PCR-confirmed SARS-CoV-2 infection

Fever is the most common symptom in the 14 days after the test, the majority of residents with a fever were in the S-based group (76% vs. 53% TP-based). The core symptoms cough and dyspnea were also most common in the S-based group compared to the TP-based group(cough 42% vs. 13% ; dyspnea 50% vs. 13%). Of all residents with confirmed COVID-19 tested based on transmission prevention, 73% eventually developed core symptoms after an average of 4.2 days (SD ±2.3). Decreased appetite was almost twice as common in the S-based group as compared to the TP-based group (64% vs. 38%). Agitation and confusion were more often reported in the TP-based group compared to the S-based group (agitated 31% vs. 6% and confusion 36% vs. 11%). Body weakness and anxiety were reported more often in the S-based group (body weakness 47% vs. 24% and anxiety 28% vs. 9%). Furthermore, the following symptoms were reported: dysphagia and somnolence, both more often in the S-based group, 25% and 31% respectively. Diarrhea, fatigue and nausea were more frequently present in the TP-based group, 24%, 27% and 18% respectively.

### Day-to-day fluctuations in temperature, 7 days prior and 14 days after testing

The mean temperature of the residents with confirmed COVID-19 was lower in the 7 days prior to the positive RT-PCR test compared to the day of the RT-PCR test or up to ∼10 days after positive RT-PCR testing (Fig.2A). About up to two days before the positive confirmed COVID-19 test an increase in temperature was observed, and on the day of the RT-PCR-test the average temperature was highest. About seven days after the positive RT-PCR test, the temperature slowly decreased again. The temperature in the S-based group was higher before testing, on the day of the RT-PCR test and after the RT-PCR test compared to residents in the TP-based group (Fig.2B).

**Figure 2.**
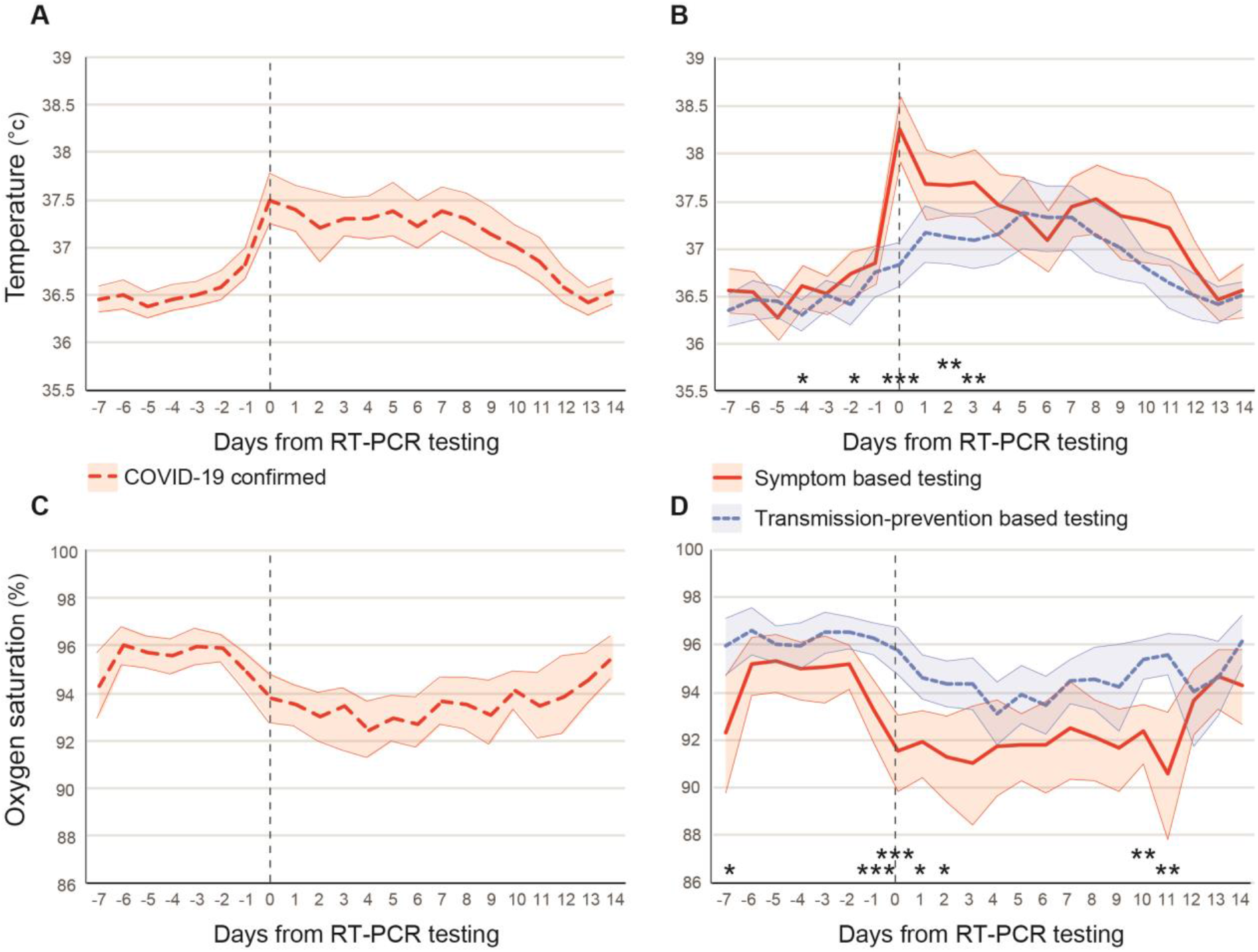
Temperature and oxygen saturation 7 days prior to the RT-PCR test until 14 days after testing for SARS-CoV-2 (T0) in LTCF residents with confirmed COVID-19 (n = 81). (A) Temperature for confirmed COVID-19. (B) Temperature for symptom based testing group (S-based n = 36) and transmission-prevention based testing group (TP-based n = 45), (C) Oxygen saturation for confirmed COVID-19. (D) Oxygen saturation for S-based and TP-based testing group. The shaded area represents the 95% confidence intervals. (*p <.05, ** p<.01, ***p<.001)

### Day-to day fluctuations in oxygen saturations, 7 days prior and 14 days after testing by residents with COVID-19± and COVID-19+

The mean oxygen saturation by the COVID-19 confirmed group was higher in the 7 days prior to the positive RT-PCR test compared to the day of the RT-PCR test or up to ∼10 days after positive RT-PCR testing. The oxygen saturation dropped about two days before the positive RT-PCR test. The oxygen saturation fluctuated until approximately 11 days after the positive confirmed COVID-19 test, after which it slowly increased again (Fig. 2C). Overall oxygen saturation was lower in the S-based group prior to testing, on the day of the RT-PCR test and after testing in comparison to the TP-based group (Fig.2D).

### Mortality in LTCF residents with confirmed COVID-19 in S-based and TP-based testing group

Of the residents with a confirmed COVID-19, 40% died within 30 days (95% confidence interval [CI], 29% to 50%) More residents in the S-based group died 56% (95% CI, 40% to 72%) compared to residents in the TP-based group 27% (95% CI, 14% to 40%) (Fig. 3). Residents with in the S-based group were 2.5 times more likely to decease within 30 days than residents in the TP-based group (adjusted hazard ratio (HR), 2.56; 95% 1.3 to 5.2; P = .010). This difference remained when adjusted for gender and age (HR of 2.4; 95% CI 1.2 to 4.9; P = .017) and when adjusted for gender, age and reduced renal function (HR of 2.2; 95% CI 1.0 to 4.6; P = .038).

**Figure 3.**
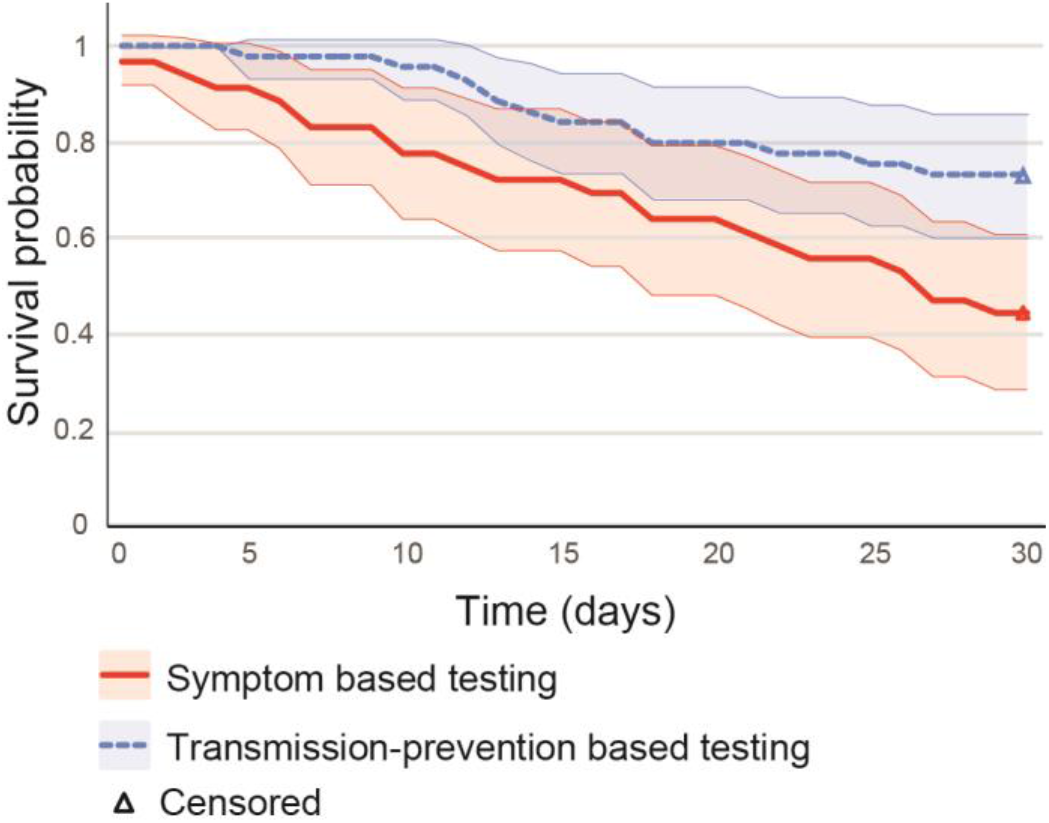
Kaplan-Meier estimates of survival for LTCF residents with COVID-19.

## Discussion

We were able to gather information about signs and symptoms about LTCF residents with confirmed COVID-19 in the week prior to the RT-PCR SARS-CoV-2 test up to two weeks after testing; we also collected data about 30 day mortality. Of the residents with confirmed COVID-19, 44% of the residents were tested based on the presence of core symptoms (S-based) and 56% of the residents were tested based on transmission prevention (TP-based). CT-values did not differ significantly between both groups.

In the 7 days prior to the test, only fever was observed as core symptom in the S-based group. We found that falling and somnolence were the most common reported other symptoms prior to the test in the TP-based group. Thus, extra attention should be paid to these symptoms since they might indicate a possible infection. This is in line with previous studies, in which symptoms such as falling and delirium were reported, but in which debuts with core symptoms were more common ^13-15^. In the 14 days following the test, apart from the core symptoms, other symptoms that were common were decreased appetite, body weakness, dysphagia, somnolence, confusion and being anxious.

Residents with confirmed COVID-19 who were tested based on transmissions prevention (TP-based) and not because of the presence of core symptoms on day of testing did develop core symptoms (fever, cough, dyspnea) in 73% of the cases, but 27% did not. To prevent transmission, we cannot base the testing policy on solely testing residents with core symptoms. The results of this study ask for repeated testing of all residents that have been in close contact with a resident with confirmed COVID-19. Furthermore all residents living together with a resident with confirmed COVID should be isolated for a quarantine period and personal protective equipment should be used in the care of these residents. Repeated testing of all care personnel of these wards is also appropriate^16^.

Our results thus support the approach for repeated testing, irrespective of symptoms, in skilled nursing facilities that has been advocated since May 2020^17^. Our study showed no difference between CT-values of S-based residents and TP based residents, similar to previous studies.^18,19,16,20^

In our study, 40% of the residents with confirmed COVID-19, died within 30-days. This percentage is higher than the 34% reported by McMichael et al.,^21^and the 26% reported by Arons et al^2^. This differences could be due to a difference in the prevalence of dementia, in our study the prevalence is much higher compared to the other studies. Moreover, we observed that residents with core symptoms on day of testing (S-based testing) were 2.5 times more likely to decease within 30 days than residents without these core symptoms on day of testing (TP-based). Thus, even though for transmission prevention all residents should be tested irrespective of symptoms, staff should still be more alert when the core symptoms are present since the mortality risk is higher for these residents as compared to residents without these symptoms. However, please note that mortality rate was still high 27% in the TP-based group and that a large part of the TP-based group did develop core symptoms after ∼4 days.

We are the first to show day-to-day fluctuations of the oxygen saturation in COVID-19 positive LTCF residents; oxygen saturation decreases approximately two days before testing in all residents who tested positive. We observed that oxygen saturation was lower for the S-based group prior to test than for the TP-based group. In addition, we observed an increase in temperature ∼ 3 days prior to test. Day-to-day fluctuations in temperature in residents with COVID-19 were described previously^9^. Yet, our day-to-day temperature measurements add to this by making a distinction between LTCF residents with and without core symptoms on day of testing. We observed that temperature for the LTCF residents without core symptoms at day of test (TP-based) also increased 2 days prior to test. These results of the daily fluctuations in temperature and oxygen saturation (shifts 1 to 2 days before the positive test) can contribute to earlier testing and earlier detection of the ‘invisible’ SARS-CoV-2 virus. If testing is done sooner, the residents can also be placed in a separated cohort earlier and the chance of possible spreading of the SARS-CoV-2 virus is smaller.

Furthermore, measuring the temperature and oxygen saturation on a daily basis is non-invasive and is independent from the ability to express symptoms. Many LTCF residents frequently have difficulty putting their symptoms into words and therefore run the risk of being missed. For example, our results demonstrated that almost all residents who were tested based on transmission prevention were living in a psychogeriatric ward and had dementia. By measuring daily changes in their temperature and oxygen saturation we may reduce the chance of transmission in this vulnerable group of LTCF residents.

This study had some limitations that must be acknowledged when interpreting results. The study was carried out within one organization and based on EHR data. Because of the use of EHR data we may have missed symptoms and also workload during the COVID-19 pandemia may have influenced symptom registration in the EHR negatively. As we used both care staff and physicians registration, this might have been somewhat overcome. Measuring the oxygen saturation and especially the temperature (tympanic and rectal) will not have been the same in every ward, but it is close to practice. A RT-PCR test has relatively low sensitivity (63-78%)^22^. Consequently we will have missed cases of COVID-19.

### Conclusions and Implications

Many LTCF residents with positive PCR did not have core symptoms when tested (fever, cough and dyspnea) but had other signs/symptoms at the day of testing and in the week before the positive test. Yet, a large part of this group, did develop these core symptoms after the testing day. Daily fluctuations in temperature and oxygen saturation can contribute to earlier detection. The results of this study underscore the importance of current testing policies that advice ample and repeated testing of all residents and personnel that are in close contact with a resident with confirmed COVID-19. This study shows that course profiles may be present. However, in order to substantiate this with more certainty, more research is needed that is prospective and longitudinal.

## Data Availability

-

## Acknowledgements

We thank the facility residents; the physicians and healthcare professionals for their ongoing efforts to provide care in the face of these challenges; M.D. Stijn Besseling and M.D. Thomas van Zijl for their help with data collection. M.D. Fleur Koene for proofreading our manuscript and input on the CT-values.

